# Effects of Varying Approaches to Lifting COVID-19 Pandemic Restrictions in the United States

**DOI:** 10.1101/2022.01.04.22268766

**Authors:** Jessica E. Galarraga, Daniel Popovsky, Kevin Delijani, Hannah Hanson, Mark Hanlon

## Abstract

**Background:** Policy approaches to lifting COVID-19 restrictions have varied significantly across the United States. An evaluation of the effects of state reopening policies on population health outcomes can inform ongoing and future pandemic responses. This study evaluates the approaches to lifting social distancing restrictions based on adherence to the Centers for Disease Control and Prevention (CDC) guidance established during the first wave of the COVID-19 pandemic.

**Methods:** We performed a retrospective study using difference-in-differences analyses to examine the effects of reopening policies on COVID-19 outcomes with risk-adjustment for population density, temporal changes, and concurrent mask policy implementation. We examined the effects of reopening policies on per capita case rates and rates of severe COVID-19 outcomes, including hospitalizations and deaths.

**Results:** Adherence to the CDC’s reopening gating metrics and phased social distancing guidelines resulted in fewer COVID-19 cases, hospitalizations, and deaths. Phase one adherent states exhibited a 50-fold reduction in daily new cases and a 3-fold reduction in daily new deaths after reopening. Phase two adherent states experienced improvements in COVID-19 outcomes after reopening, while non-adherent states had a resurgence of worsening outcomes after lifting restrictions.

**Conclusions:** Our study findings indicate that adherence to the CDC’s reopening guidance after implementing social distancing restrictions during the COVID-19 pandemic substantially prevents new cases, hospitalizations, and deaths. Following a stepwise reopening strategy and ensuring a sustained decline in case rates and test positivity rates before lifting restrictions can mitigate on a large scale the negative effects of a pandemic on population health outcomes.

## INTRODUCTION

The novel coronavirus disease-2019 (COVID-19) caused by the severe acute respiratory syndrome coronavirus 2 (SARS-CoV)-2 led to the deadliest, fastest-moving pandemic since the Spanish flu from over a century ago. As of January 4, 2022, over 290 million COVID-19 cases have been reported globally and 5.5 million individuals have died from the disease.^1^ The United States (U.S.) is among the countries most affected by the pandemic, accounting for approximately 20% of all COVID-19 confirmed cases while representing 4% of the global population.^1,2^ Since the onset of the pandemic, the U.S. has had a predominately decentralized public health response, with state governments determining when and how to lift COVID-19 restrictions.^3–6^ The heterogeneity of policy approaches observed in the U.S. offers an opportunity to examine its effects on epidemiological outcomes and improve our understanding of effective reopening strategies during a pandemic.

Several studies from the U.S. and other countries have demonstrated a well-established inverse relationship between social distancing measures and COVID-19 case rates.^7–12^ For example, prior research by Courtemache et al. examining the effects of mitigation policies found that shelter-in-place orders and closures of restaurants, gyms, bars and entertainment venues mitigated the spread of COVID-19 after 16 to 20 days.^12^ However, there remains to be an evaluation of the approaches to reopening and its effects on COVID-19 outcomes to guide the lifting of restrictions during a pandemic. Prior work has been limited to examining raw trends during reopening phases by state without testing the effects of the differences in reopening procedures.^13^

After the deployment of vaccines and the subsequent, albeit temporary, improvement of case rates in June 2021, U.S. states, the District of Columbia (DC), and Puerto Rico lifted remaining restrictions towards full reopening. However, the surge driven by the Omicron (B.1.1.529) variant, a COVID-19 virus for which early findings demonstrate a remarkably high transmissibility rate with an ability to evade natural and vaccine-induced immunity, has led to the re-implementation of social distancing restrictions to mitigate spread.^15, 16^ As of December 22, 2021, multiple European nations, including Italy, Germany, France, Portugal, Finland, Scotland, Wales, Northern Ireland, and the Netherlands, have re-implemented social distancing measures.^17^ Additionally, multiple U.S. states and Puerto Rico have implemented new gathering restrictions based on vaccination status in response to the Omicron variant.^14, 18^

The long-term trajectory of the ongoing COVID-19 pandemic and the potential for future pandemics secondary to new infectious agents underscore the importance of understanding the efficacy of reopening strategies based on when and how social distancing restrictions are lifted. This research examines the effects of different policy approaches to lifting COVID-19 restrictions on epidemiological outcomes in the U.S., including case, hospitalization, and death rates. In this study, we account for population densities, temporal changes, and the presence of mask mandates and use per capita measures to evaluate the effects of varying approaches to reopening. U.S. states were classified based on their adherence to the Centers for Disease Control and Prevention’s (CDC) phased framework for reopening and guidance on gating metrics to proceed with lifting social distancing restrictions.^19–21^

## METHODS

We performed a retrospective study of state-level policy and epidemiological COVID-19 data in the United States from March 8 through July 4, 2020. We focus on the period of the first pandemic wave, when there was the least within state heterogeneity in reopening policies and the CDC had provided gating metrics as a guidance for states to take a measured approach to reopening. We included data on all 50 states, the District of Columbia, and Puerto Rico.

### Epidemiological Data

COVID-19 epidemiological data was collected from the Center for Systems Science and Engineering (CSSE) at Johns Hopkins University.^1^ The COVID Tracking Project has assessed the overall quality of the data for each state and territory included in the CSSE database based on the completeness of data reporting.^22^ The majority of states and territories (93%) had a grade of “A” or “B” in data quality, while 7% received either a “C” or “D.” We collected data on new daily cases, hospitalizations, and deaths for each state over a four-month period starting from the initial date daily COVID-19 outcomes became available in the CSSE database, March 8, 2020.

Outcome measures included daily new case rates to examine the spread of COVID-19 across the population as well as daily new hospitalizations and deaths to examine the burden of COVID-19 disease. All measures were calculated per 1 million population. The pre-pandemic population denominator for each U.S. state was based on U.S. Census data from 2019.^2^ Consistent with the approach used by the CDC, 7-day rolling averages of the outcome measures were used to account for spurious outliers.^23^

### State Policy Data

We developed a database documenting reopening policies implemented in response to the COVID-19 pandemic for each of the 50 U.S. states, the District of Columbia, and Puerto Rico. We compiled data from gubernatorial and Department of Health resources on state policy directives for each phase of reopening during the study period and documented whether or not they adhered to two domains of the CDC’s guidance: the gating criteria for reopening and the phased framework for social distancing measures to be lifted during phases one and two of reopening.^24^ The gating criteria to support the progression of lifting COVID-19 restrictions included (1) a decreasing 14-day trend in newly identified COVID-19 cases, (2) a decreasing 14-day trend in the COVID-19 test positivity rate, and (3) sufficient hospital capacity defined as inpatient and intensive care unit bed occupancy less than 80%.^18-20^ We did not include the number of visits for symptoms of influenza-like illness from the CDC’s gating criteria since the study period was not within influenza season, the period when this data is captured.

We defined CDC adherence as meeting the gating criteria at the time of reopening and being either consistent with or stricter than the social distancing guidance for the corresponding phase of reopening. We defined non-adherence as not meeting gating criteria and being more permissive in the lifting of social distancing restrictions than the CDC guidance. We also classified states by whether they adhered to only the gating metrics or only the phased framework for social distancing to assess the individual efficacy of the two domains of reopening guidance.

### Statistical Analysis

The effects of CDC guidance adherence with phase one and two reopening on daily new COVID-19 case, hospitalization, and death rates per million population were examined using difference-in-differences (DiD) analyses. Analyses were risk-adjusted for population density, defined as population per square mile, the presence of concurrently implemented mask policies, weekly fixed effects to account for temporal variation common to both the case and control groups, and state fixed effects to address unobserved, non-time-varying differences between states, with standard errors clustered on the state level.^25^

For all statistical analyses, p<0.05 (2-sided test) was used as the significance level. SPSS version 27 (IBM Corporation, Armonk, NY) was used for database construction activities and SAS version 9.4 (SAS Institute, Inc., Cary, NC) for statistical analyses. This study was approved by the Institutional Review Board as non-human subjects research.

## RESULTS

### Sample Characteristics

We studied COVID-19 epidemiological outcomes across 50 U.S. states, the District of Columbia, and Puerto Rico, with a total of 6,188 daily observations in the study sample. Across all states, the 7-day rolling average case rate at the time of lifting COVID-19 restrictions was lower among states whose policies adhered to the CDC’s guidance for phase one reopening but similar for phase two reopening. The mean difference between adherent and non-adherent states in the case rate per 1 million population was 23.6 (90.9 vs. 67.3) for phase one reopening and 1.8 (58.4 vs. 56.6) for phase two reopening (Table 1).

**Table 1.** COVID-19 Reopening Policies and Corresponding 7-Day Rolling Average Case Rates Per 1 Million Population at Time of Implementation.

|  |  |
| --- | --- |
| **Number of States/Territories** | 52 |
| **Phase One Reopening<sup>†</sup>** |  |
| Adherence (mean, [SE]) | 90.9 (2.5) |
| Non-Adherence (mean, [SE]) | 67.3 (1.1) |
| **Phase Two Reopening<sup>†</sup>** |  |
| Adherence (mean, [SE]) | 58.4 (0.8) |
| Non-Adherence (mean, [SE]) | 56.6 (0.8) |

<sup>†</sup>Adherence is defined as reopening according to the CDC guidance on reopening phases when gating metrics have been met, and non-adherence is defined as lifting restrictions more loosely than the CDC guidance and not meeting gating metrics. COVID-19 = 2019 novel coronavirus

### CDC Guidance Adherence of Reopening Plans

States whose reopening policies were adherent to the CDC guidance had a reduced rate of daily new cases, hospitalizations, and deaths per 1 million population that was statistically significant. For phase one reopening, CDC adherence led to a 94 per million (95% CI: −104.6, −83.3) fewer daily new cases, 34.6 per million (95% CI: −38.3, −30.9) fewer daily new hospitalizations, and 3.6 per million (95% CI: −4.2, −2.9) fewer daily new deaths. CDC adherence for phase two reopening policies demonstrated similar findings, with adherence resulting in 94.2 (95% CI: −104.1, −84.4), 13.6 (95% CI: −17.9, −9.3), and 2.5 (95% CI: −3.3, −1.8) fewer daily new cases, hospitalizations, and deaths per million population, respectively (Table 2). Pre-post rate differences for each reopening phase and outcome measure are provided in Table 2.

**Table 2.** Difference-in-differences (DiD) Analysis of COVID-19 Outcomes per Million Population based on State Adherence to CDC Reopening Guidance

|  | Pre-Post Rate Difference<sup>‡</sup><br>(95% CI) | DiD Estimate<sup>‡</sup><br>(95% CI) |
| --- | --- | --- |
| Phase One Reopening<sup>†</sup> |  |  |
| Daily New Cases |  |  |
| Adherence | -95.9 (-109.7, -82.1)** | -94.0 (-104.6, -83.3)** |
| Non-Adherence | -1.9 (-13.9, 10.1) |  |
| Daily New Hospitalizations |  |  |
| Adherence | -23.8 (-28.1, -19.5)** | -34.6 (-38.3, -30.9)** |
| Non-Adherence | 10.8 (7.2, 14.5)** |  |
| Daily New Deaths |  |  |
| Adherence | -4.9 (-5.8, -4.2)** | -3.6 (-4.2, -2.9)** |
| Non-Adherence | -1.4 (-2.1, -0.7)** |  |
| Phase Two Reopening<sup>†</sup> |  |  |
| Daily New Cases |  |  |
| Adherence | -27.4 (-37.8, -16.9)** | -94.2 (-104.1, -84.4)** |
| Non-Adherence | 66.9 (56.7, 77.0)** |  |
| Daily New Hospitalizations |  |  |
| Adherence | -2.4 (-2.1, 6.8) | -13.6 (-17.9, -9.3)** |
| Non-Adherence | 15.9 (11.3, 20.6)** |  |
| Daily New Deaths |  |  |
| Adherence | -0.8 (-1.5, -0.03)* | -2.5 (-3.3, -1.8)** |
| Non-Adherence | 1.7 (1.0, 2.5)** |  |

\* Difference has a statistically significant p-value < .05, \*\* p-value ≤ 0.0001
<sup>†</sup> Adherence is defined as reopening according to the CDC guidance on reopening phases when gating metrics have been met, and non-adherence is defined as lifting restrictions more loosely than the CDC guidance and not meeting gating metrics.
<sup>‡</sup> Linear probability model risk-adjusted for mask policy implementation, population density, and state/territory and week fixed effects. Daily new case, hospitalization, and death rates are based on the 7-day rolling average per 1 million population.
COVID-19 = 2019 novel coronavirus; CDC = Centers for Disease Control and Prevention; 95% CI = 95% Confidence Interval

Temporal trends in daily new cases among the study groups demonstrate that phase one adherent states maintained a declining trend in cases after week 9, which corresponds to the median week of phase one reopening across the states. On the other hand, non-adherent states exhibited a relatively stable trend during the same time period. (Figure 1) Similarly, phase 2 adherent states maintained a declining trend after week 13, which corresponds to the median week of phase two reopening, while non-adherent states experienced an increasing trend after phase 2 reopening. (Figure 2) When examining individually the trends in cases among states that adhered to only one of the domains of the CDC guidance, the improvement in cases was more pronounced with adherence to the social distancing measures for phase one, while the improvement in cases was more pronounced with adherence to gating metrics for phase two.

**Figure 1.**
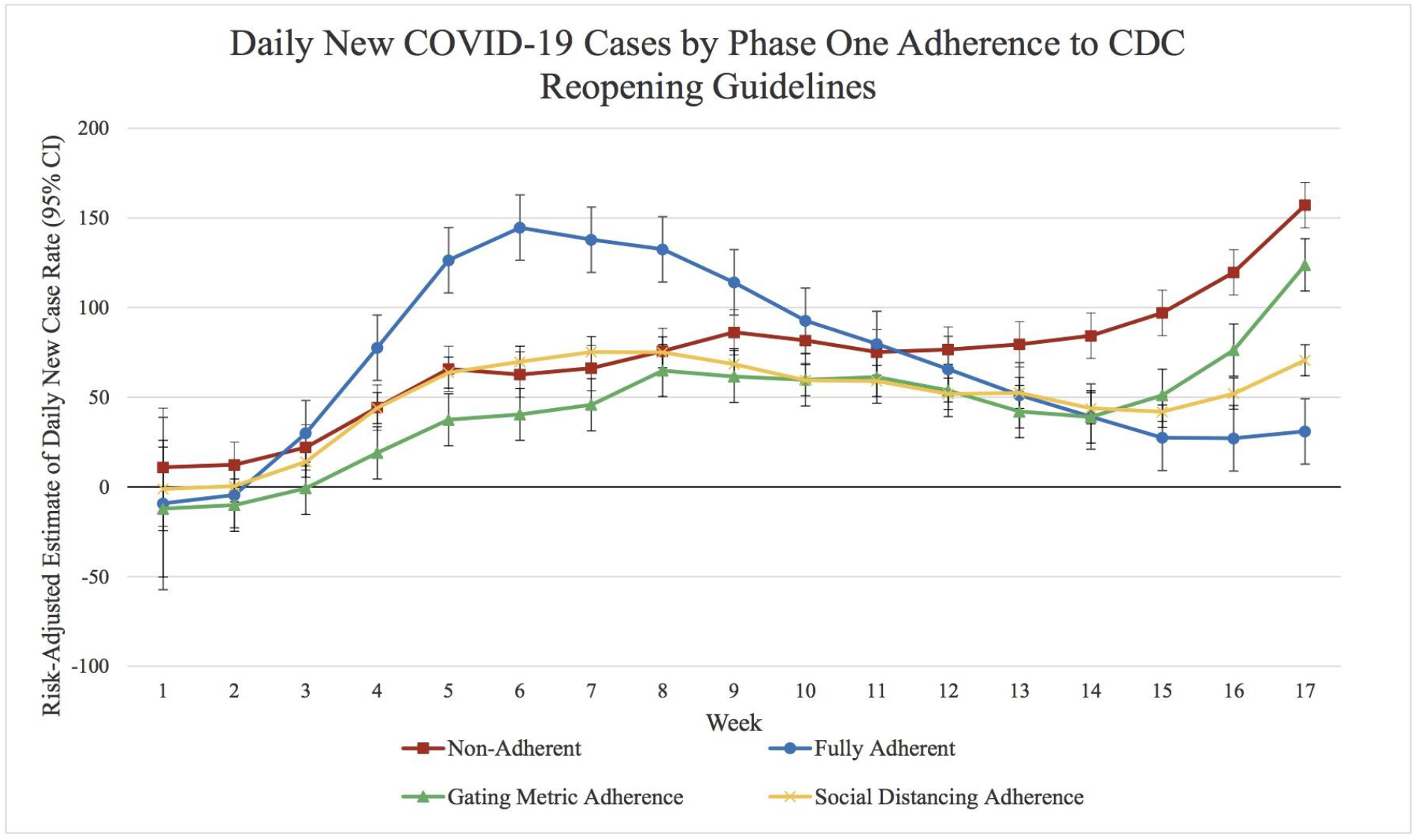
Times series analysis of weekly levels of risk-adjusted COVID-19 case rates by level of adherence to phase one reopening guidelines by the Centers for Disease Control and Prevention (CDC), including (1) non-adherence, (2) adherence to gating metrics only, (3) adherence to social distancing only, or (4) fully adherent with adherence to both gating metrics and social distancing. Week one corresponds to March 8, 2020 through March 14, 2020. Case rates represents the 7-day rolling average of daily new cases per 1 million population and were adjusted for population density, mask policy implementation, and both state and week fixed effects, with standard errors clustered by state. Vertical lines show 95% confidence intervals (CIs).

**Figure 2.**
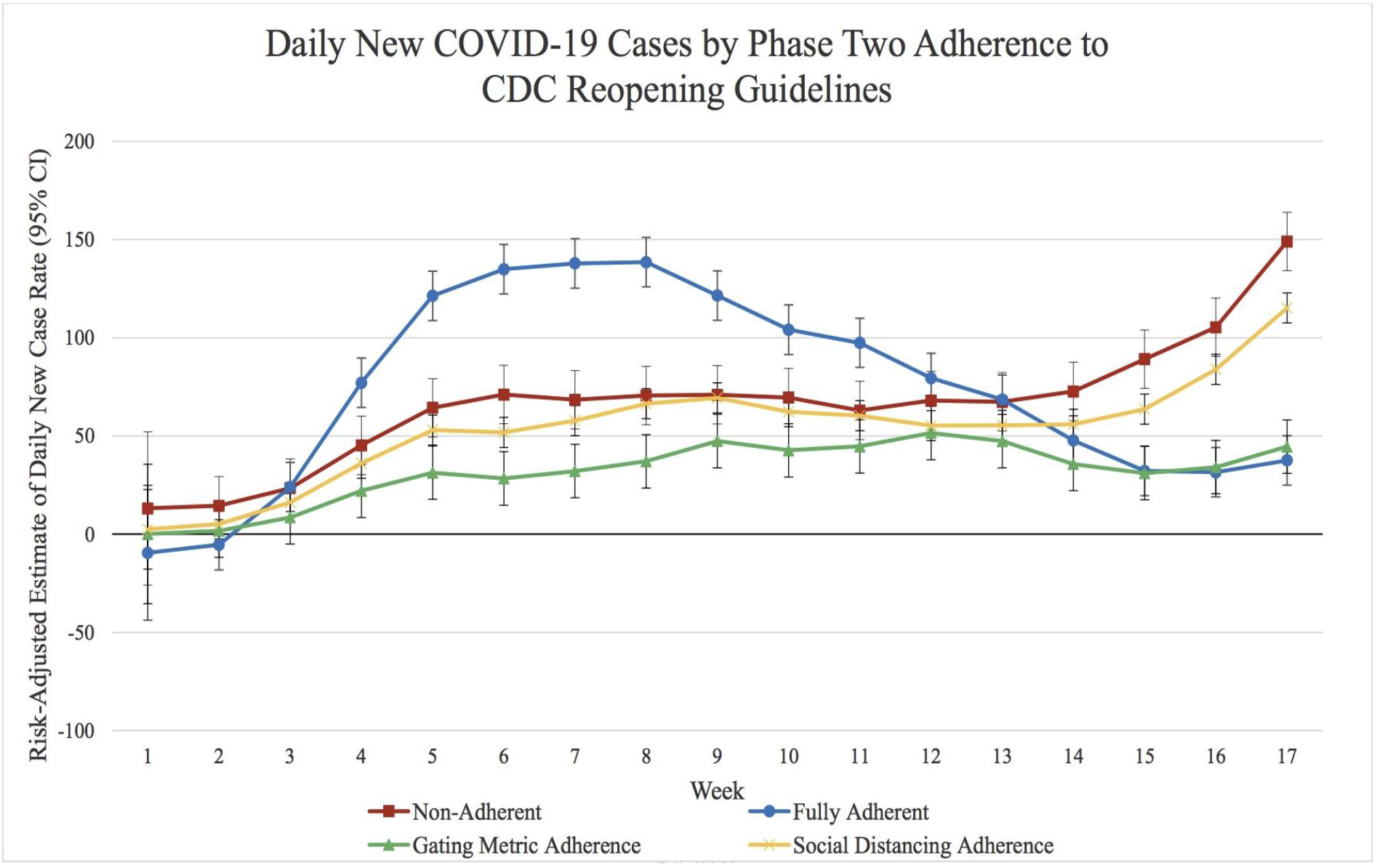
Times series analysis of weekly levels ofrisk-adjusted COVID-19 case rates by level ofadherence to phase two reopening guidelines by the Centers for Disease Control and Prevention (CDC), including (1) non-adherence, (2) adherence to gating metrics only, (3) adherence to social distancing only, or (4) fully adherent with adherence to both gating metrics and social distancing. Week one corresponds to March 8, 2020 through March 14, 2020. Case rates represents the 7-day rolling average of daily new cases per 1 million population and were adjusted for population density, mask policy implementation, and both state and week fixed effects, with standard errors clustered by state. Vertical lines show 95% confidence intervals (CIs).

## DISCUSSION

The heterogeneity of state reopening policies during the COVID-19 pandemic provides a unique opportunity to explore the effects of varying approaches to lifting pandemic restrictions on health outcomes. This investigation is the first to conduct a risk-adjusted assessment of the reopening approaches implemented across the U.S. on epidemiological measures. Our study provides evidence on the efficacy of using a data-driven, phased approach when lifting pandemic restrictions, which is based on a sustained decline in new cases and test positivity with sufficient hospital capacity prior to lifting social distancing measures.

The findings of this study indicate that adherence to the CDC’s reopening guidance after implementing social distancing restrictions is effective in preventing substantial rates of daily new COVID-19 cases, hospitalizations, and deaths. Phase one adherent states exhibited a 50-fold reduction in daily new cases and 3-fold reduction in daily new deaths compared to those that were non-adherent. Phase one adherence was also associated with reduced hospitalizations while non-adherent states experienced increases in daily new hospitalizations. Phase two adherence was also associated with reduced daily new case, hospitalization, and death rates, while the non-adherent group experienced increases in all three outcomes after reopening.

Based on the mean U.S. state population of 5.9 million, our study findings on phase one adherence alone would translate into 555 cases, 204 hospitalizations, and 21 deaths that are prevented on a daily basis across an average populated state such as Wisconsin. On the national level, our study findings translate into 1,182 fewer deaths per day with phase one reopening adherence and 821 fewer deaths per day with phase two reopening adherence based on the U.S. population.

The points of diverging trends in daily new cases among adherent versus non-adherent states are consistent with when the majority of states progressed into their next phase of reopening during our study period, with daily new case rates increasing among non-adherent states while adherent states maintained declining case rates after reopening. Additionally, case rates with phase one reopening, when restrictions are first lifted following stay-at-home orders, appear to be more sensitive to adherence to social distancing measures, while case rates with phase two reopening are more sensitive to adherence to gating metrics.

There are several limitations to this study. First, although we focused on a period of the pandemic with the least within state heterogeneity in policy responses, there was still county-level variation within some U.S. states in reopening policies during the study period which may lead to heterogeneity within the comparison groups. This limitation, however, would underestimate rather than overestimate the effects of adherence and lessen our ability to detect significant differences between the study groups. Despite this limitation, our study findings detected substantial disparities in outcomes by adherence category on the state level. Another study limitation is that the adherent group included some states that were stricter than the CDC guidance in the phased framework for social distancing and therefore the documented effects may be partly driven by restrictions stricter than the tested CDC guidelines. Further, the study period is limited to the first wave of the pandemic in the U.S. and the magnitude of the effects of reopening strategies on epidemiological trends with the presence of vaccination and differences in the transmissibility of COVID-19 may vary.

## CONCLUSIONS

The variation in reopening policies across U.S. states during the COVID-19 pandemic has provided new insights on epidemiological outcomes associated with the approach to lifting social distancing restrictions. Following a stepwise reopening strategy and ensuring a sustained decline in case rates and test positivity rates before lifting restrictions can mitigate on a large scale the negative effects of a pandemic on population health outcomes.

## Data Availability

All data produced in the present study are available upon reasonable request to the authors

